# Severe Sarcoidosis Demonstrates Heterogenous Immune Dysregulation Across Different Peripheral Immune Cell Types: A Single-Center Single Cell Multi-omic Analysis

**DOI:** 10.1101/2024.09.18.24313886

**Authors:** Kai Huang, Christen Vagts, Christian Ascoli, Yue Huang, Nadera J. Sweiss, Patricia W. Finn, David L. Perkins

**Affiliations:** University of Illinois at Chicago; University of New Mexico

## Abstract

Sarcoidosis provides unique management challenges due to variable presentation and disease course. In this single-center analysis, we assess transcriptomic and epigenomic differentiators between severe and mild cases of sarcoidosis using peripheral blood. We showcase differences across multiple cell types that highlight decreased immunoregulation and increased inflammation in severe sarcoidosis. We provide an overall framework for the interactions between immune cells in severe sarcoidosis as a basis for future therapeutic research.

**Visual Abstract:** 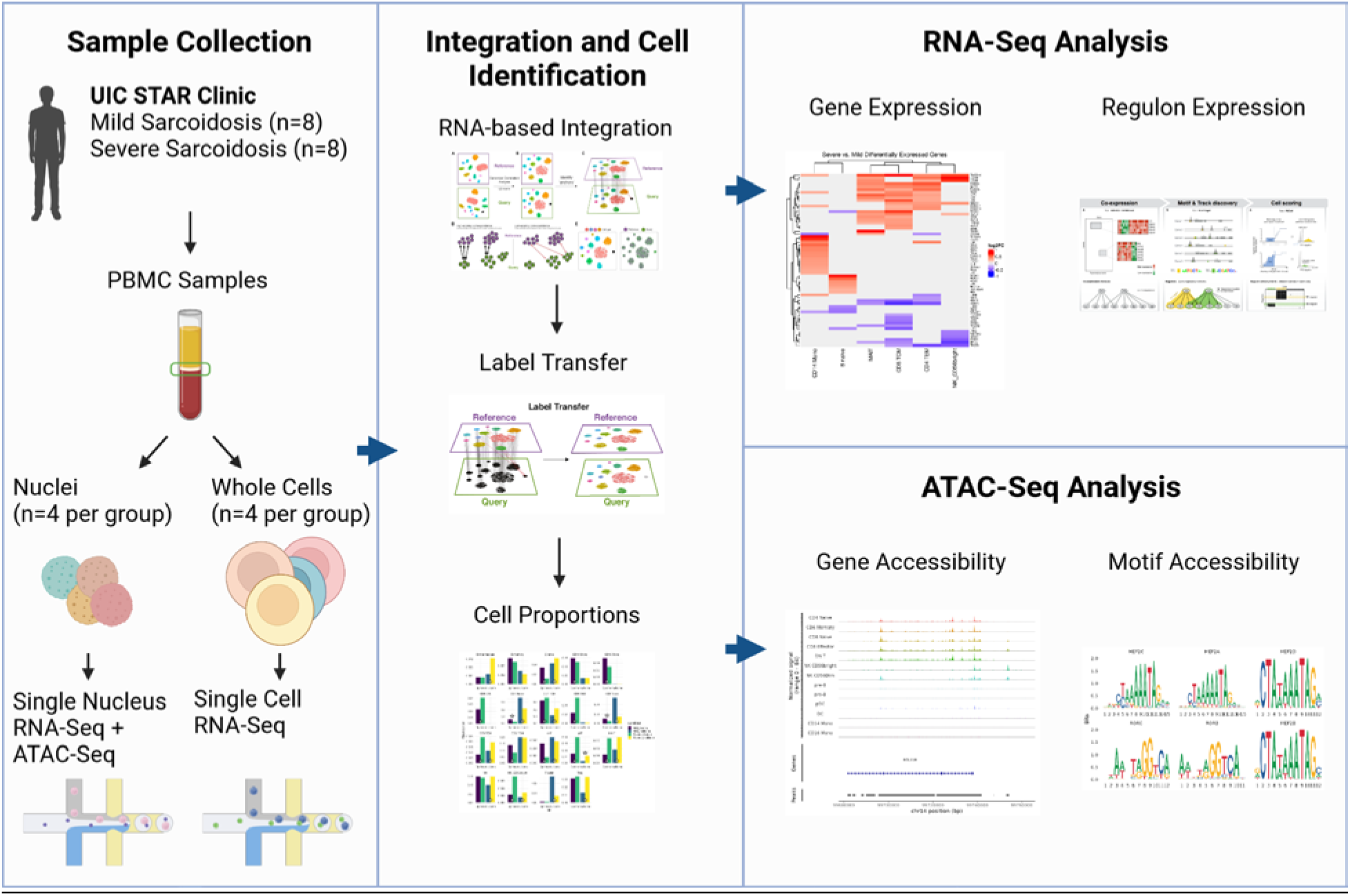

## Intro

Sarcoidosis is an immune mediated multiorgan disease characterized by granulomatous inflammation. Pulmonary involvement is the most common disease manifestation(BAUGHMAN et al., 2001; Judson et al., 2012) and can be highly variable, ranging from asymptomatic imaging abnormalities to progressive symptomatic disease resulting in diminished quality life and respiratory failure. Respiratory failure as the leading cause of disease related mortality(Swigris et al., 2011), however cardiac, neurologic, and ocular involvement are additional contributors to disease related morbidity and mortality (Baughman et al., 2010; Judson, 2015; Morimoto et al., 2008). While two thirds of patients may experience spontaneous resolution within 2-5 years(Valeyre et al., 2014), the natural course of disease is highly variable and difficult to predict. Identified risk factors for persistent disease include multiple organs involved, the presence of neurologic or cardiac involvement, and those with a greater severity of dyspnea (Doubková et al., 2015; Rodrigues et al., 2011). The risk of morbidity and mortality, as indicated by more severe disease, is heavily weighted in the decision to initiate treatment with immunosuppression (Baughman et al., 2017). Severe disease can necessitate long term immunosuppressive therapy with steroids as well as additional disease modifying anti-sarcoidosis drugs (DMASDs) which carry additional risks (Ungprasert et al., 2019). However, understanding of the mechanisms that result in severe disease is lacking.

Existing literature suggests numerous cell types and subpopulations contribute to sarcoidosis. Our prior works on peripheral blood mononuclear cells (PBMC) indicate that aberrant proinflammatory, exhaustive, and anergic pathways are associated with features of sarcoidosis disease severity and outcome (Ascoli et al., 2022; Schott et al., 2019). However, transcriptomic and proteomic studies have relied on bulk sequencing techniques which have been unable to elucidate cell-specific pathways to further define disease mechanisms. Single-cell RNA sequencing (scRNA-seq) is a newer modality that provides an unbiased evaluation of transcriptomic mechanisms underlying each cell population and has provided insight into the unique cell types in which sarcoidosis-specific pathways are dysregulated (Garman et al., 2020; Krausgruber et al., 2023; Liao et al., 2021). These studies implicate dysfunctional macrophages, monocytes, and T cells among peripheral blood, bronchoalveolar lavage (BAL), and skin compartments, which aligns with bulk sequencing studies. Liao et al. specifically employed a multi-omic technique through use of scRNA-seq and bulk ATAC-seq (assay of transposase accessible chromatin) to identify epigenetic variation among BAL macrophages of individuals with progressive sarcoidosis (Liao et al., 2021). Epigenetics, which is understudied in sarcoidosis, may provide insight into the heterogeneous presentation and outcomes of sarcoidosis despite significant gene overlap.

We utilize single cell RNA-sequencing along with single nuclei multiome sequencing which allows combined ATAC-seq and RNA-seq to broadly profile the immune transcriptome and epigenome of PBMCs in individuals with severe and non-severe sarcoidosis. It has been shown that well-expressed marker genes are highly correlated between these modalities and that utilizing expression data from whole cells and nuclei can aid consistent cell identification (Sandoval et al., 2023). Thus, the RNA-seq data from both kits were integrated and analyzed together. From this multiomic dataset, we performed differential expression, differential accessibility, and transcription factor activity analysis. While there is no unified definition of severity in sarcoidosis, we considered subjects with the highest risk of morbidity and mortality to be have the most severe disease (i.e. those with cardiac, ocular, or central neurologic involvement, as well as those with pulmonary disease characterized by forced vital capacity <50%). We hypothesize that severe sarcoidosis is characterized by a distinct transcriptomic and epigenetic immunologic signature that will elucidate key mechanisms of sarcoidosis progression and provide new strategies for disease management.

## Methods

### Recruitment and Sample Collection

Study approval was obtained through the University of Illinois at Chicago (UIC) IRB Ethics Review Committee, Approval #2016-0063. Subjects with biopsy-proven sarcoidosis, diagnosed in accordance with ATS/ERS/WASOG criteria,(Costabel & Hunninghake, 1999) were recruited. All subjects were older than 18 years of age and received their sarcoidosis care in the Bernie Mac Sarcoidosis Translational Advanced Research (STAR) Center at UIC. Subject clinical data was also extracted from their medical charts at the time of sample collection. Any patients with neurologic, cardiac, or ocular sarcoidosis as well as those with forced vital capacity (FVC) less than 40% of predicted values are considered to have severe disease.(Sweiss et al., 2010)

Blood samples were collected from patients visiting the STAR Center for routine follow-up. PBMCs were extracted from fresh blood using density gradient centrifugation at 400g with Ficoll-Paque PLUS. Extracted PBMCs were stored in FBS with 10% DMSO in liquid nitrogen for single cell preparation.

### Single Cell RNA-Seq

Cryopreserved PBMCs were processed for sequencing via the 10X Genomics Chromium Single Cell 3’ V3.1 reagents with dual indexing. To prepare the cells for single cell partitioning on the 10X Chromium device, cells were thawed at 37C then immediately serially diluted in warm sterile RPMI (Fisher# 22400089) with HEPEs and without antibiotics. The PBMCs were then transferred into HBSS with 0.04% BSA and processed according to the 10X Genomics protocol with a targeted cell recovery of ∼10,000 cells per sample.

### Single Cell Multiome-Seq

Cryopreserved PBMCs were processed for sequencing via the 10X Genomics Chromium Single Cell Multiome ATAC + Gene Expression Kit. Cells were thawed via the same protocol as the single cell 3’ kit. After transferring cells to HBSS the cells were lysed to extract nuclei according to the 10X Genomics nuclei extraction protocol. The resulting nuclei were immediately processed according to the multiome kit’s protocol with a targeted cell recovery of ∼10,000 nuclei per sample.

### Sequencing and Data Processing

Libraries were assessed for quality via the Agilent Bioanalyzer. Subsequently, libraries were pooled and sequenced using one NovaSeq S2 flowcell for all RNA-seq libraries (including both 3’ and multiome kits) and one NovaSeq S2 flowcell for all ATAC-seq libraries (only from multiome kits) targeting 25,000 reads per cell for RNA-seq and 50,000 reads per cell for ATAC-seq. Raw sequencing files were trimmed via fastp.(Chen et al., 2018) RNA-seq libraries were then aligned and quantified using alevin with an intron-aware hg38 reference.(Srivastava et al., 2019) ATAC-seq libraries were quantified using Cell Ranger ARC (10X Genomics) with the hg38 reference. Processed files were imported into R using the tximeta and Signac packages for RNA and ATAC libraries respectively.(Love et al., 2020; Stuart et al., 2021)

### Filtering and Integration

Each sample was imported into a Seurat object and preprocessed via the sctransform pipeline within the Seurat package.(Hafemeister & Satija, 2019) RNA counts for the 3’ libraries consisted of only the exon counts while RNA counts for the multiome libraries consisted of both introns and exons due to the significantly higher proportion of intronic reads in nuclei. Data from all assays were filtered by the following inclusion criteria: RNA count greater than 1000 and total genes detected between 200 and 3000. Additionally, 3’ kit cells with more than 15% of reads attributed to mitochondrial transcripts and multiome kit nuclei with more than 20% of reads attributed to mitochondrial transcripts were filtered out. The mitochondrial filter for nuclei was set higher due to higher proportions of such reads in nuclei.(Basile et al., 2021) Libraries generated from the multiome kit were also filtered by including only nuclei with ATAC-seq counts between 2000 and 70,000. Only cells and nuclei passing all filterers were retained for further analysis.

The dataset was then integrated using Seurat’s reciprocal PCA method with sctransform normalization on the RNA counts. After integration, the integrated data was used to transfer cell type labels from a multiomodal CITE-seq reference dataset onto our dataset.(Hao et al., 2021) Doublets were identified and visualized via DoubletFinder and removed from the dataset.(Stoeckius et al., 2018)

### Dimension Reduction

The data was then split to further process the multiome and 3’ kits independently to leverage strengths of each modality. For the 3’ data, the CITE-seq reference’s supervised PCA (sPCA) dimension reduction scheme was integrated with our data to generate a new sPCA that included the variability within our sarcoidosis samples. The integrated sPCA was used to generate a new UMAP dimension reduction to improve the visualization and clustering of this data. Since the multiome data is based on nuclei but the CITE-seq reference is based on cells, sPCA transfer to the multiome data was not sufficient for visualization. Instead, the multiome data was further processed by first computing an integrated latent semantic indexing (LSI) dimension reduction using the ATAC-seq data. Subsequently, the ATAC-seq based LSI dimension reduction was combined with a PCA computed from the multiome data’s RNA-seq data to conduct a weighted nearest neighbor analysis (WNN).(Hao et al., 2021) A UMAP dimension reduction was generated using WNN to facilitate visualization of the multimode dataset.

### Differential Expression and Differential Accessibility

Differential expression analysis was completed using the sctransform normalized RNA counts for all samples via the FindMarkers function in Seurat using the MAST algorithm.(Finak et al., 2015) Gene ontology analysis was applied to the DEGs using the topGO package with FDR correction.(Alexa & Rahnenfuhrer, 2020) Transcription factor and gene regulatory network inference was applied to the RNA data across the entire dataset using pySCENIC implemented in the VSN-Pipelines workflow.(Aibar et al., 2017; Flerin et al., 2021; Van de Sande et al., 2020) Both SCENIC analyses modes were utilized, one using known and inferred binding motifs from cisTarget, and one using known ENCODE ChIP-seq tracks.

Single-cell level transcription factor activity inferred by pySCENIC was then imported into Seurat and used for differentially expressed regulon (DER) analysis via FindMarkers with the Wilcoxin Rank-Sum test and the log2FC threshold set to 0.005 due to regulon activity having much finer fluctuations than differential gene expression.

The ATAC-seq data from multiome samples was further analysed via motif enrichment and differential accessibility. Motif accessibility was quantified with the JASPAR2022 motif database and the chromVAR R package.(Castro-Mondragon et al., 2022; Schep et al., 2017) After quantification, differentially enriched motifs (DEM) were calculated for each cell type using Seurat’s FindMarkers function with the Wilcoxin Rank-Sum test with FDR correction. Differentially accessible genes (DAGs) were calculated directly from the ATAC-seq data using the FindMarkers function with a logistical regression with FDR correction and regressing out the number of fragments overlapping peaks in each individual cell. Differential peaks were then given a gene identity by assigning each peak to the gene closest to it on the chromosome. All differential analyses were performed only on samples with at least 5 cells of the cell type being assessed.

For each differential comparison (DEGs, motif-based DERs, track-based DERs, DEMs, DAGs), cell types were ranked by how many genes/motifs/peaks were detected in each modality with the highest rank assigned to the cell type with the highest count of differential results. Ribosomal or mitochondria transcripts were excluded from the ranking calculation. Ranks were then averaged across all five modalities. Two separate PCAs were calculated from the log2 fold-change values of differential elements from transcriptome and epigenome respectively for visualization.

## Results

### Demographics

Our aim in this study was to expand upon the bulk transcriptome findings in the previous sarcoidosis severity study and apply single cell level analyses to further elucidate severity dependent changes in immunological activity and pinpoint cell types that drive the various facets of immune dysfunction found in sarcoidosis. A total of 16 patients with sarcoidosis were recruited for this study, with 8 cases of severe sarcoidosis and 8 cases of mild sarcoidosis. Patient demographic details are shown in Table I.

**Table I:** Patient demographics for single cell experiments split by severity and single cell library kit. 3’ indicates the normal 3’ single cell RNA-seq kit while multiome indicates the 10X multiome RNA-seq plus ATAC-seq kit.

|  | Non Severe Disease |  | Severe Disease |  | Overall |  |
| --- | --- | --- | --- | --- | --- | --- |
|  | 3'<br>(N=4) | multiome<br>(N=4) | 3'<br>(N=4) | multiome<br>(N=4) | 3'<br>(N=8) | multiome<br>(N=8) |
| **Age** |  |  |  |  |  |  |
| Mean (SD) | 58.5 (6.51) | 53.4 (8.42) | 62.4 (6.23) | 52.2 (2.85) | 60.5 (6.26) | 52.8 (5.85) |
| Median [Min, Max] | 60.3 [49.7, 63.7] | 56.5 [41.1, 59.7] | 65.2 [53.1, 66.1] | 52.2 [48.8, 55.8] | 63.4 [49.7, 66.1] | 53.9 [41.1, 59.7] |
| **Sex** |  |  |  |  |  |  |
| Female | 2 (50.0%) | 3 (75.0%) | 2 (50.0%) | 3 (75.0%) | 4 (50.0%) | 6 (75.0%) |
| Male | 2 (50.0%) | 1 (25.0%) | 2 (50.0%) | 1 (25.0%) | 4 (50.0%) | 2 (25.0%) |
| **Ethnicity** |  |  |  |  |  |  |
| Black | 2 (50.0%) | 3 (75.0%) | 1 (25.0%) | 2 (50.0%) | 3 (37.5%) | 5 (62.5%) |
| White | 2 (50.0%) | 1 (25.0%) | 2 (50.0%) | 2 (50.0%) | 4 (50.0%) | 3 (37.5%) |
| Hispanic | 0 (0%) | 0 (0%) | 1 (25.0%) | 0 (0%) | 1 (12.5%) | 0 (0%) |
| **BMI** |  |  |  |  |  |  |
| Mean (SD) | 33.5 (4.94) | 31.7 (7.35) | 36.4 (5.77) | 32.6 (8.79) | 35.0 (5.21) | 32.1 (7.52) |
| Median [Min, Max] | 32.3 [29.0, 40.5] | 30.5 [24.0, 41.7] | 34.6 [32.0, 44.6] | 31.9 [22.6, 44.0] | 32.8 [29.0, 44.6] | 31.4 [22.6, 44.0] |
| **No Treatment** |  |  |  |  |  |  |
| No | 4 (100%) | 4 (100%) | 3 (75.0%) | 4 (100%) | 7 (87.5%) | 8 (100%) |
| Yes | 0 (0%) | 0 (0%) | 1 (25.0%) | 0 (0%) | 1 (12.5%) | 0 (0%) |
| **Corticosteroids** |  |  |  |  |  |  |
| No | 0 (0%) | 1 (25.0%) | 1 (25.0%) | 1 (25.0%) | 1 (12.5%) | 2 (25.0%) |
| Yes | 4 (100%) | 3 (75.0%) | 3 (75.0%) | 3 (75.0%) | 7 (87.5%) | 6 (75.0%) |
| **Antimetab** |  |  |  |  |  |  |
| No | 3 (75.0%) | 3 (75.0%) | 2 (50.0%) | 2 (50.0%) | 5 (62.5%) | 5 (62.5%) |
| Yes | 1 (25.0%) | 1 (25.0%) | 2 (50.0%) | 2 (50.0%) | 3 (37.5%) | 3 (37.5%) |
| **HCQ** |  |  |  |  |  |  |
| No | 4 (100%) | 2 (50.0%) | 4 (100%) | 4 (100%) | 8 (100%) | 6 (75.0%) |
| Yes | 0 (0%) | 2 (50.0%) | 0 (0%) | 0 (0%) | 0 (0%) | 2 (25.0%) |
| **TNF** |  |  |  |  |  |  |
| No | 4 (100%) | 4 (100%) | 4 (100%) | 2 (50.0%) | 8 (100%) | 6 (75.0%) |
| Yes | 0 (0%) | 0 (0%) | 0 (0%) | 2 (50.0%) | 0 (0%) | 2 (25.0%) |
| **IVIG** |  |  |  |  |  |  |
| No | 3 (75.0%) | 4 (100%) | 4 (100%) | 4 (100%) | 7 (87.5%) | 8 (100%) |
| Yes | 1 (25.0%) | 0 (0%) | 0 (0%) | 0 (0%) | 1 (12.5%) | 0 (0%) |

### Integration and Preprocessing Results from Combined Analysis of 3’ and Multiome Kits Reveal Changes in Cell Type Abundance Related to Severity

Prefiltering of the raw single cell transcriptomes resulted in a total of 125,500 cells passing filter. After doublet detection and removing detected doublets, a total of 92,731 cells were retained for analysis. The 3’ RNA-seq kit yielded 44,973 cells while the multiome kit yielded 47,758 cells. After integration and label transfer, all cell types from the reference were detected in our dataset. A total of 19 cell types of 30 in the reference had total cell counts higher than 150 cells and were used for further analysis. The proportions of all cell types are shown in Figure 1. Naïve B cells showed a decrease in abundance in severe patients sequenced on the 3’ kit but increased in abundance in severe patients on the multiome kit. All other cell types either only showed significant differences in one kit or had trends in the same direction in both kits. Some notable trends corroborated by both kits included a severity dependent increase in abundance of both CD4+ and CD8+ T central memory cells (TCMs) as well as gamma-delta T cells (gdTs), membrane-associated invariant T cells (MAITs), and CD56bright natural killer cells (NKs). CD8+ naïve T cells showed a significant decrease in both kits in severe patients. Some differences in cell type abundance between the two kits were observed and expected due to the different extraction methods used in each kit. Notably, CD16+ monocytes and CD4+ cytotoxic T lymphocytes (CTLs) had fewer than 10 cells detected in the multiome kit with 421 and 1364 cells detected respectively in the 3’ kit. Additionally, platelets had only 43 cells detected from the 3’ kit versus 1044 cells detected from the multiome kit.

**Figure 1:**
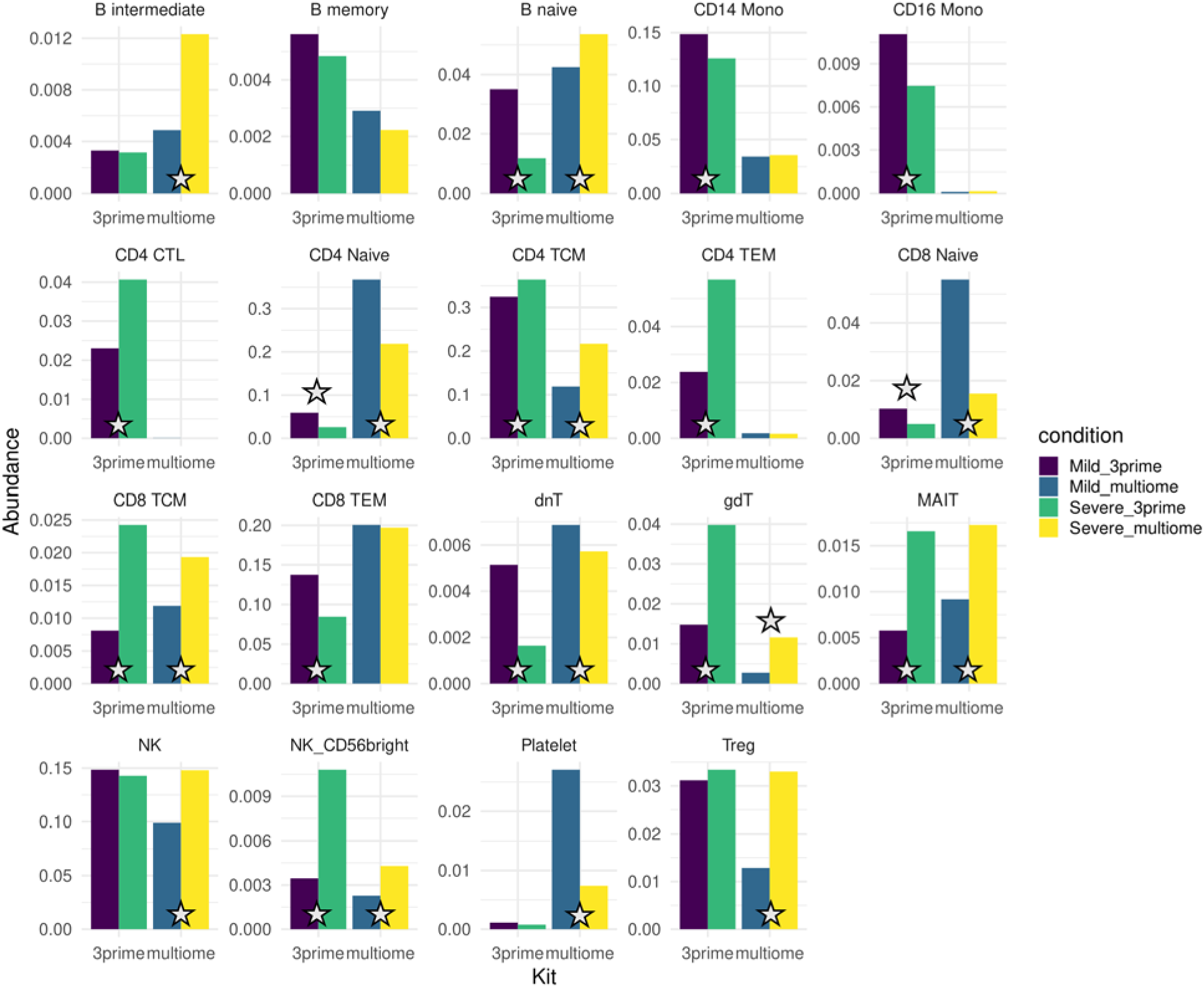
Severe sarcoidosis exhibits differences in the proportion of various cell type. Abundances that were significantly different between mild and severe disease samples from the same sequencing kit are marked with a star (FDR-adjusted p<.05). Due to differences in extraction technique, abundance differences between kits were not assessed for significance. Cell type name abbreviations: cytotoxic T lymphocyte (CTL), monocytes (mono), T central memory (TCM), T effector memory (TEM), double negative T (dnT), gamma-delta T (gdT), membrane-associated invariant T (MAIT), natural killer (NK), regulatory T (Treg).

Visualization of the integrated data via a UMAP shows no remaining sample or disease severity batch effects after integration. Limited separation was observed between the multiome and 3’ kits due to the kits using nuclei and cells respectively. However, this did not affect cell type determination (Figure 2). These findings show agreement with Further analysis of UMAPs generated from the 3’ kit alone or the multiome kit alone showed consistent separation between different cell types (Figure 3 and 4).

**Figure 2:**
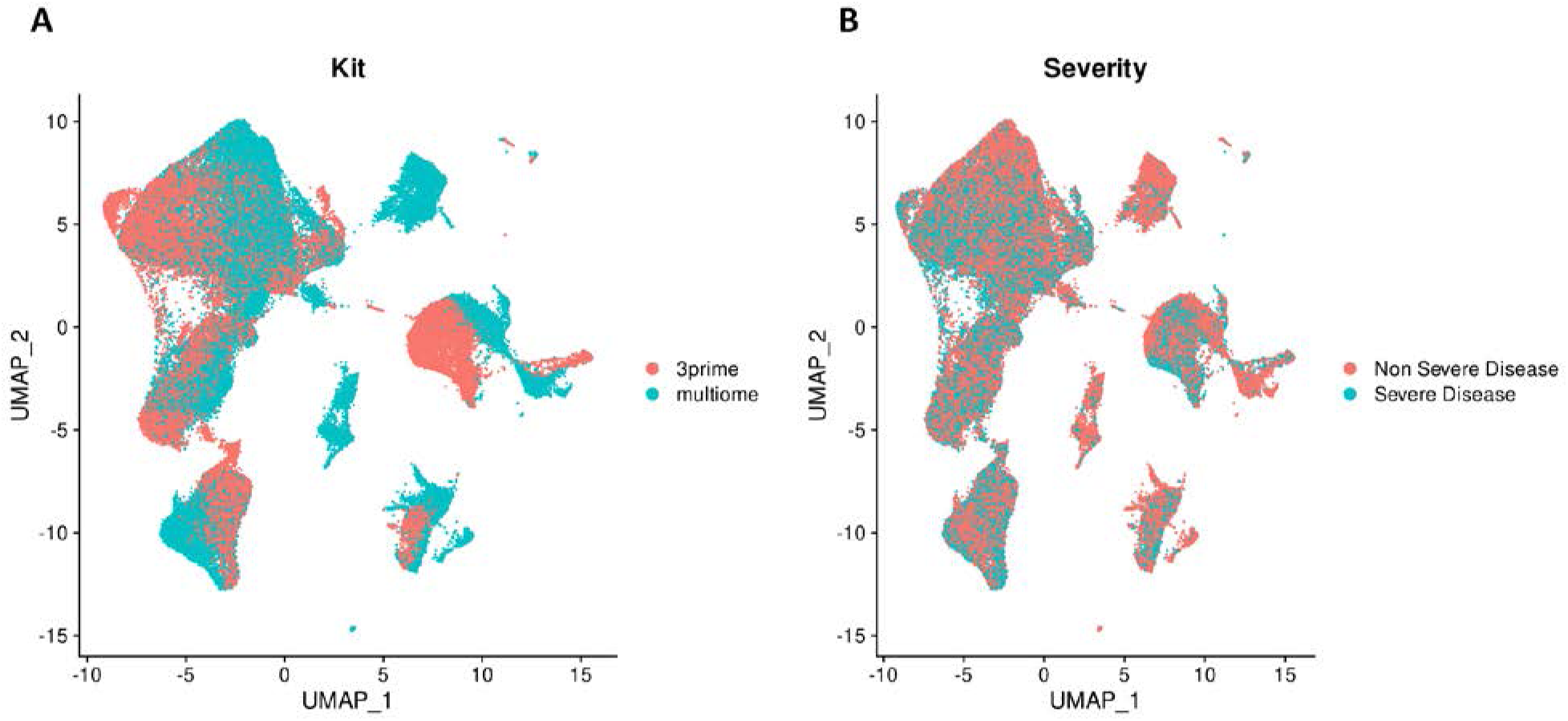
UMAP distribution of cells based on sequencing kit and disease severity. **A.** A UMAP visualization of cells that passed filtering from both kits. Some separation between cells from each kit it observed due to differences in cell recovery and kit cell isolation method. **B.** A UMAP visualization of cells passing filter separated by patient severity. Cells from both groups show overlap indicating successful integration.

**Figure 3:**
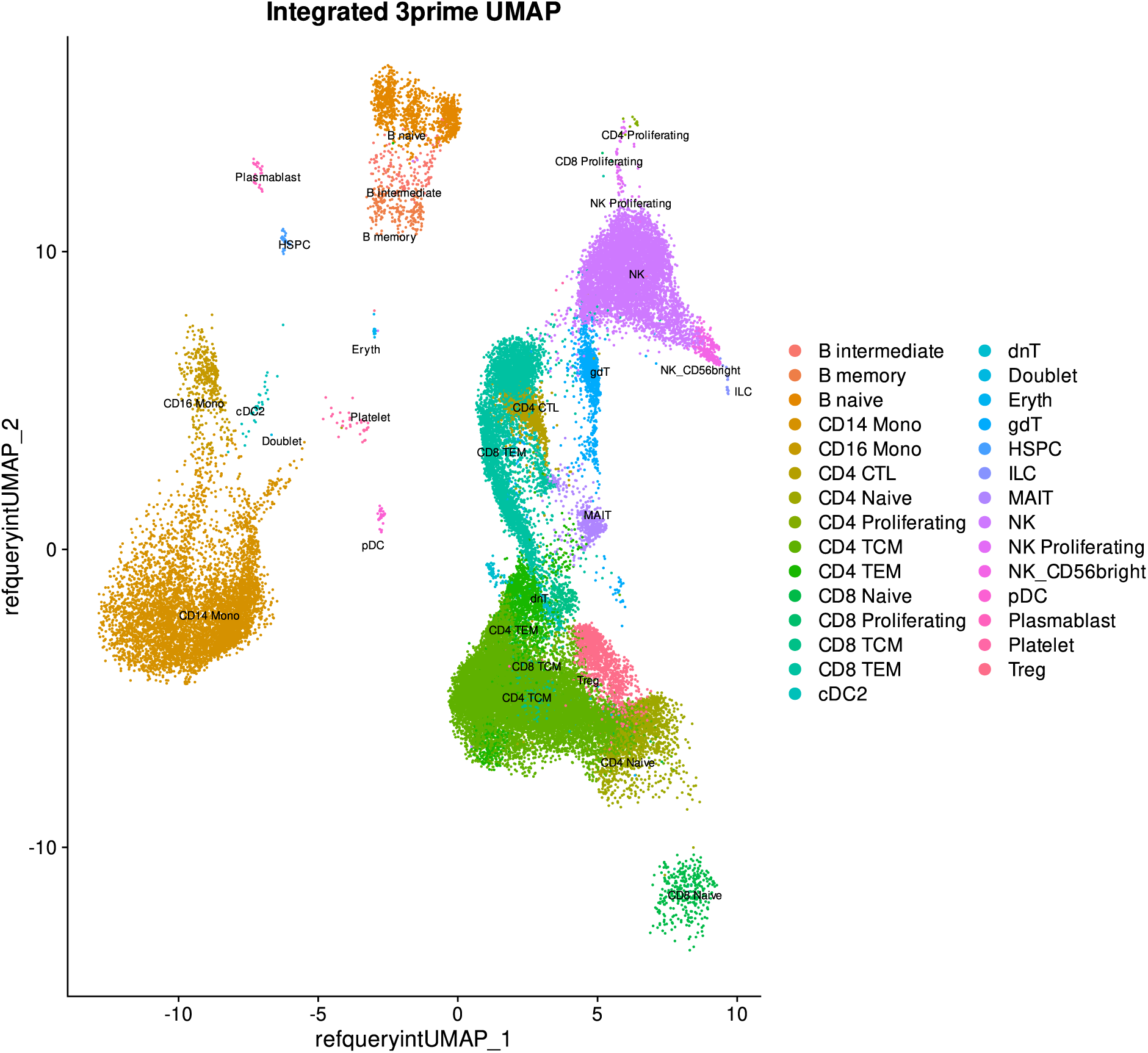
Cell identifications inferred from multi-modal CITE-seq and RNA-seq data for 3’ RNA-seq samples. The UMAP projection was calculated by integrating the reference UMAP and our 3’ data UMAP to generate a combined plot utilizing information from both.

**Figure 4:**
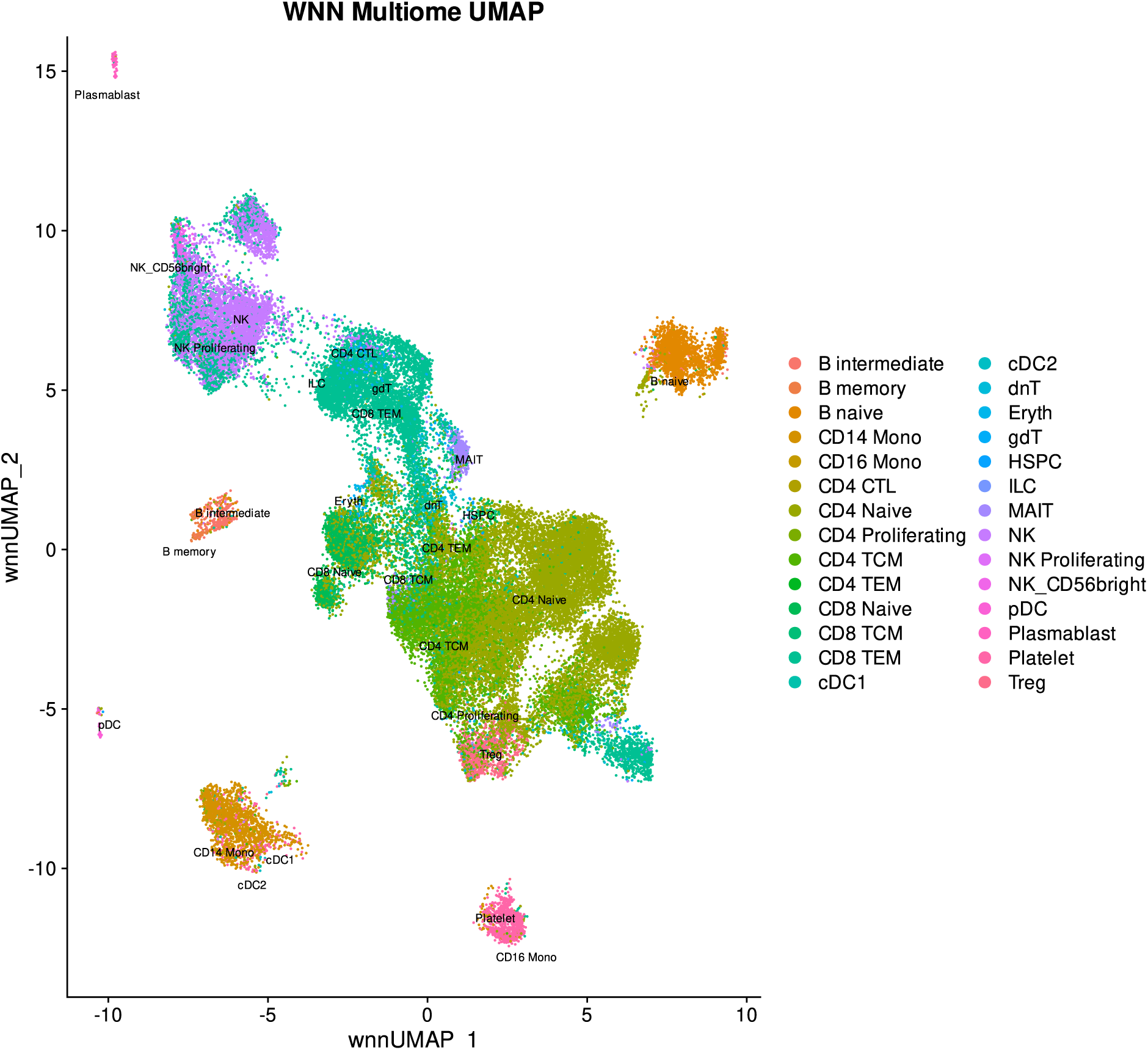
Cell identifications inferred from multi-modal CITE-seq and RNA-seq data for multiome samples. This UMAP is calculated from RNA-seq and ATAC-seq data in the multiome samples without including the UMAP from the reference since it did not contain ATAC-seq data. RNA-seq and ATAC-seq modalities were integrated via a weighted nearest neighbor (WNN) algorithm.

### Gene Ontology Analysis of DEGs in Severe Cases Highlights Changes in Metabolism and Immune Response

DEG analysis on each of the 19 cell types that passed filtering with enough cells remaining for analysis yielded a median of 58 DEGs per cell type with a minimum of 3 DEGs from double negative T cells (dnTs) and maximum of 377 DEGs from CD8+ TCMs. Due to low cell counts, CD16+ monocytes and CD4+ CTL cells did not include any multiome samples. Top GO terms in genes with increased expression in severe sarcoidosis included various immune system activation terms such as innate immune response, T cell activation, and immune response. Specifically, these terms were observed in NK cells, CD14+ monocytes, gdTs, and CD4+ T effector memory cells (TEMs). Additionally, naïve B cells showed enrichment of upregulated DEGs for B cell activation. GO terms indicative of cell metabolic activity and protein synthesis such as cytoplasmic translation and ribosomal small subunit assembly were also observed. Cell types exhibiting these GO terms in their upregulated DEGs included CD8 naïve cells, CD8 TCMs, CD56 NK cells, MAITs and CD4 CTLs. Notably, many of the cell types with metabolic and immune related upregulated GO terms are cells involved in direct immune response.

GO terms for genes with decreased expression in severe sarcoidosis contrasted with the GO terms from DEGs with increased expression. Cell types exhibiting immunoregulatory roles or hybrid roles in immune response showed top GO terms for their downregulated DEGs related to translation, immune response, and cellular activation. CD4 naïve cells, Tregs, and gdTs all had downregulated DEG GO terms for cytoplasmic translation and peptide metabolism. Additionally, CD8 TCMs, CD4 CTLs and MAIT cells showed downregulated DEG GO terms for immune response related functions and lymphocyte activation. B naïve cells also showed downregulated DEG GO terms for ribosomal subunit assembly, while B intermediate and B memory cells showed terms for gas transport.

### Cell Type Ranking and Grouping Isolates Key Cell Types with High Severity-Related Variance

The PCAs constructed using our transcriptomic and epigenomic analyses respectively revealed several cell types with high variability due to differences between mild and severe groups. In the transcriptomic analysis, naïve B cells, platelets, CD8+ TCMs, MAITs, and NK CD56bright cells were the most distinct while naïve B cells and CD14+ monocytes were most distinct in the epigenomic PCA (Figure 5).

**Figure 5:**
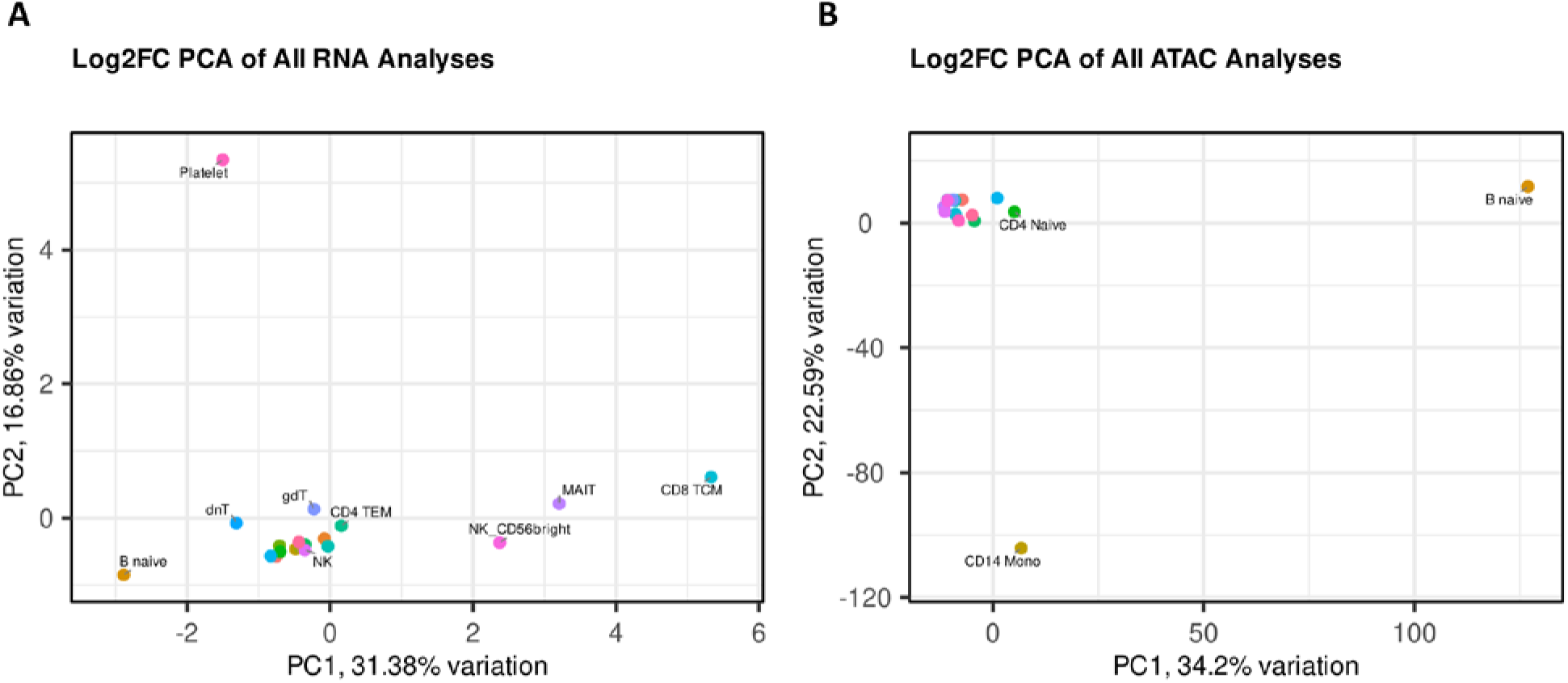
PCAs constructed from differential expression results across all modalities reveal cell types with the highest variability in our dataset. **A.** PCA constructed from all RNA-based analyses which included the log2FC values for all significant genes from differential gene expression as well as transcription factor activities from both motif and track-based SCENIC inference methods. **B.** PCA constructed from all ATAC-based analyses which included log2FC values for all significant genes from differential motif enrichment and differential accessibility.

The overall cell type ranking which included average ranks from both modalities for cells that were present in both modalities showed similar trends. The highest variance cell types were CD56bright NK cells, naïve B cells, CD8+ TCMs, MAITs, CD4+ TEMs, and CD14+ monocytes. This group largely contains cell types tied to rapid immune response except for naïve B cells. Interestingly, Tregs and CD16+ monocytes which are cell types associated with immunoregulatory roles are near the bottom of these rankings, showing minimal changes between mild and severe disease.

### DEGs within High Variance Cell Types Have Contrasting Alterations in Expression of Immune Response Related Genes

Among the six high variance cell types we identified, CD56bright NK cells, CD4+ TEMs, CD8+ TCMs, and MAITs shared similar patterns of increased expression in various proinflammatory genes in severe sarcoidosis patients. This includes increased GZMA and GZMK expression in MAIT, CD4+ TEMs, and CD56bright NKs, as well as IL7R, IL32, CD52 and LTB in MAIT, CD4+ TEMs and CD8+ TCMs.(Stelzer et al., 2016) The immunoregulator ANXA1 is also increased in expression in CD4+ TCMs, CD8+TEMs, and CD56bright NKs.(Gavins & Hickey, 2012) This proinflammatory pattern is further reinforced by expression of various S100 genes. These cell types also show decreased expression of various components of immune mediators within the JAK-STAT and PI3K/AKT and MAPK pathways such as STAT4, AKT3, and PIK3R1.(Stelzer et al., 2016) Other genes with notably decreased expression include the immune regulators FOXP1 and KLF12.(Brown et al., 2016; Guerau-de-Arellano et al., 2022; Zhang et al., 2021). Additionally, MAITs had highly increased expression of KLRB1 which has been tied to Th17 cell activation.(Ramesh et al., 2014) CD14+ monocytes had more distinct expression patterns with a mix of proinflammatory and immunoregulatory factors. These cells showed increased expression of various proinflammatory factors including CXCL2/8, CCL3/44, S100A12, FGL2, CD52, FCN1 and IL1B. This is complemented by increased expression of immunoregulatory and immunosuppressive genes such as NFKBIA, EGR1, CD74, and NFKBIZ.(Figueiredo et al., 2018; Trizzino et al., 2021). Additionally, the lncRNA MALAT1 which has multiple immunological functions was also decreased in expression.(Amodio et al., 2018; Cai et al., 2020; Masoumi et al., 2019)

Naïve B cells also had a distinct expression pattern with increased expression of B cell activation and signaling genes such as BANK1, BLK, FCRL1, LYN, PAX5, and BCL11A.(Brodie et al., 2018; DeLuca et al., 2021; Hernández et al., 2021; Medvedovic et al., 2011; Yin et al., 2019) Additionally, increased expression of HLA-DQA1 is also observed. The naïve B cells exhibited decreased expression of several immunoregulatory genes including RACK1 and KLF2.(Wittner & Schuh, 2021; Yao et al., 2013) MALAT1 was also underexpressed in these cells.

Two additional notable genes were TMSB4X and ACTB.(Cooke et al., 2013; Severa et al., 2019) TMSB4X is associated with pulmonary sclerosis (Severa et al., 2019) and was detected with increased expression in severe patients in B memory, CD14+ Monocytes, CD4+ TEMs, CD8+TCMs, gdTs, MAITs, CD56bright NKs, and platelets. ACTB is associated with severe sarcoidosis and fibrosis (Cooke et al., 2013), and was found to be increased in expression in CD4+ TEMs, CD8+ TCMs, gdTs, MAITs, NKs, platelets, and Tregs. Results for top genes of immunological interest are shown in Figure 6.

**Figure 6:**
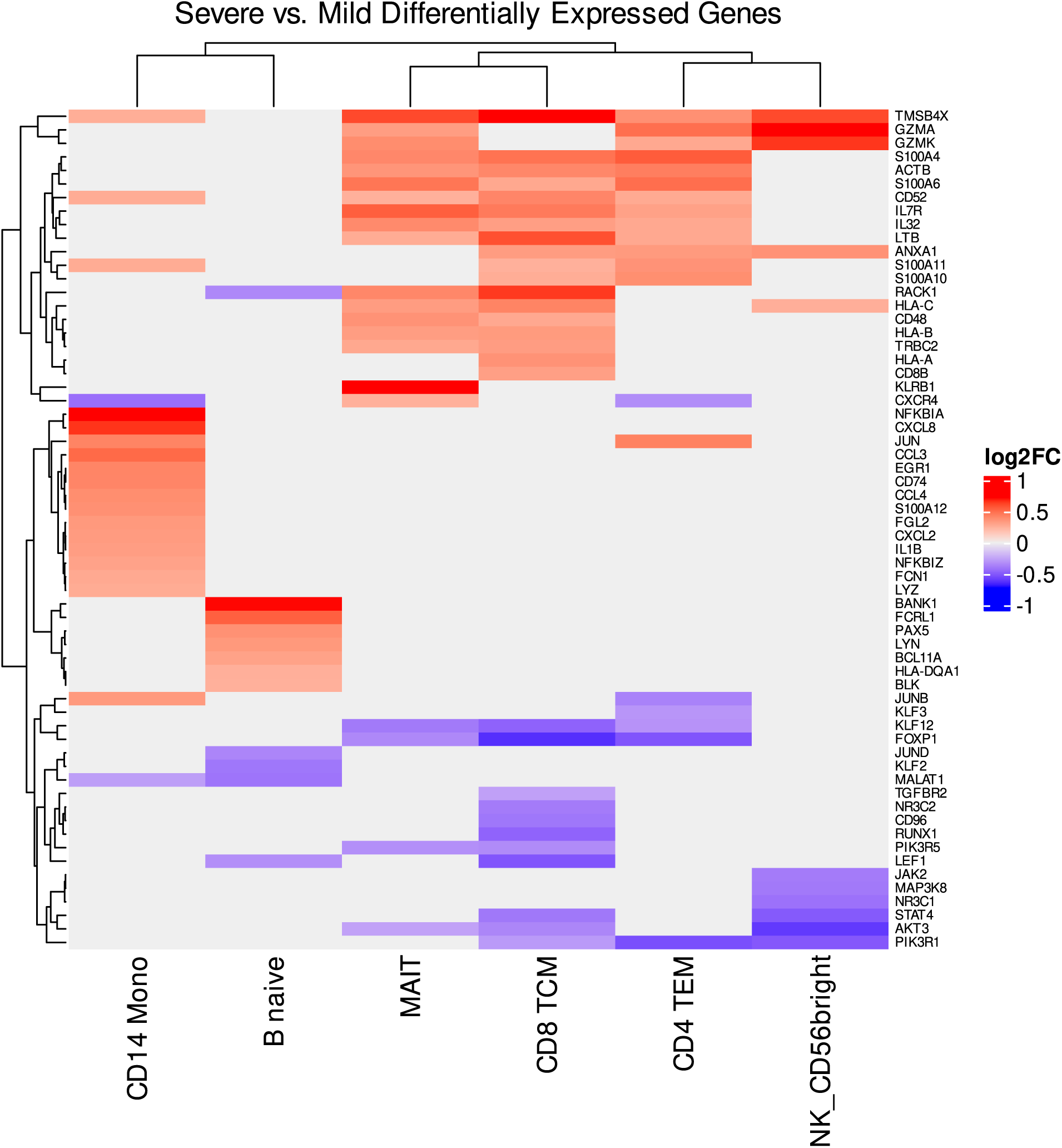
Selected DEGs with immunological functions in the high variance cell types. The heatmap plots log2FC values for significant DEGs (FDR-adjusted p-value<0.05) comparing severe cases to mild cases.

### SCENIC Regulon Activity Shows Severity-Dependent Alterations in Key Immune Response Regulators

Regulon analysis via SCENIC yielded a median of 79 DERs for binding motif-based analysis and a median of 14 DERs for track-based analysis. Within the motif analysis, CD56bright NKs, MAITs, CD8+ TCMs, CD14+ monocytes and CD4+ TEMs had similar DER profiles with a largely proinflammatory pattern (Figure 7). These cell types showed increased activity of regulons related to effector immune cell differentiation such as BATF and RORC which induces Th17 differentiation among other functions.(Guntermann et al., 2017; Shetty et al., 2022) Proinflammatory regulons such as IRF3, JUND, and FOS also showed increased activity.(Moon et al., 2017; Yanai et al., 2018) JUN, JUNB, and GATA2 also showed increased activity but excluding CD4+ TEMs, with GATA2 also having no significant change in CD14+ monocytes.(Takai et al., 2021) The cells also show increased activity of immunoregulatory and suppressive regulons including FOXP3, and IRF2.(Sheikh & Utzschneider, 2022; Ziegler, 2007)

**Figure 7:**
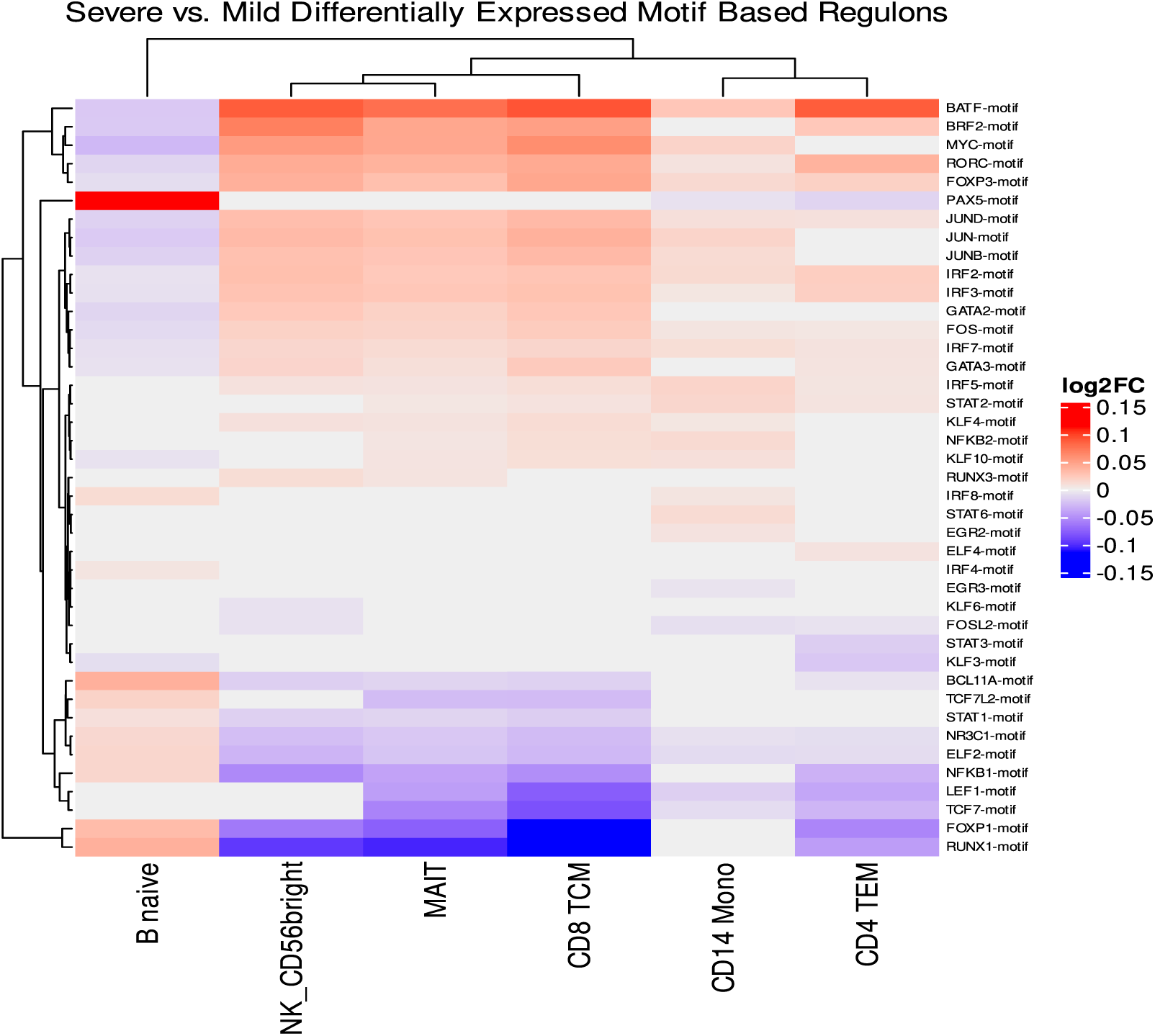
Differential expression of motif-based regulons in severe vs. mild disease for top ranked cell types. Regulon activity was inferred via SCENIC and differentially expressed regulons with immunological functions were selected. Only regulons with differential activity are shown.

The same five cell types also showed similar decreased activity in several regulators of immune cell differentiation and maturation such as ELF2 along with decreased activity of the glucocorticoid receptor NR3C1.(Guan et al., 2017; Xiao et al., 2019) BCL11A, and NFKB1 were also decreased in activity in these cell types except for CD14+ monocytes.(Tuijnenburg et al., 2018; Yin et al., 2019) The immunoregulators RUNX1 and FOXP1 also show decreased activity in this group except for CD14+ monocytes while TCF7 and LEF1 show decreased activity except in CD56bright NK cells.(Brown et al., 2016; Korinfskaya et al., 2021; Ma et al., 2022; Shan et al., 2021)

The naïve B cells showed a largely opposing trend in regulon activity to the previous five cell types, including deactivation of BATF, MYC, RORC, FOS, GATA2, JUND, JUN, and JUNB. This is contrasted by activation of differentiation and maturation regulons including PAX5, BCL11A, ELF2, and NFKB1 alongside activation of RUNX1 and FOXP1.

The track-based regulon results showcase similar trends (Figure 8). Excluding naïve B cells, the five other cell types show increased activity of both inflammatory and immunoregulatory regulons including MYC, GATA2, IRF2, RFX5, and TBX21 with MYC and GATA2 showing insignificant changes in CD4+ TEMs. Regulons with decreasing activity again involved many immune regulators such as BCL11A, STAT3, TCF7, FOSL1, and FOSL2 although only TCF7 was significant across all five cell types. Additionally, the glucocorticoid receptor NR3C1 significantly decreased in activity across all five cell types. The track-based results for naïve B cells also continue to oppose those of the over 5 cells with decreased activity in MYC, GATA2, IRF2, and TBX21. This coincided with increased activity of PAX5, BCL11A, and NR3C1.

**Figure 8:**
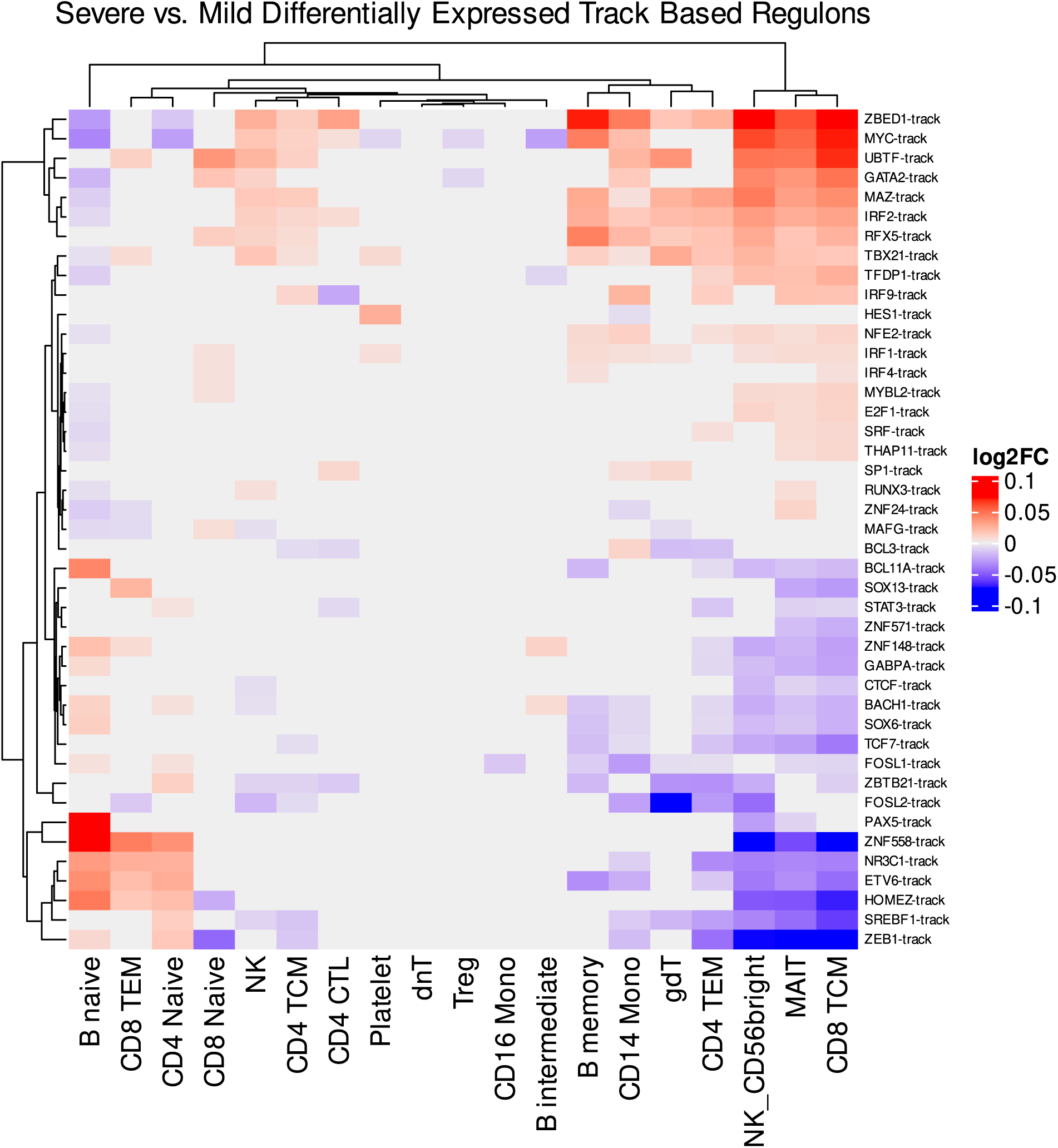
Differentially expressed track-based regulons for all cell types. Regulon activity was inferred via SCENIC. All differential regulons across the dataset are shown since the track-based method yields fewer regulons than the motif-based method.

### WNN ATAC-Seq Confirms Previous Findings and Reveals Potential Skewing Towards a Regulatory B Cell Lineage

Since CD56bright NKs and CD4+ TEMs were absent from the ATAC-seq data due to low detection, we focused on the four top ranked cell types that were still present: naïve B cells, MAITs, CD8+ TCMs, and CD14+ monocytes.

Motif enrichment analysis of MAITs and CD8+ TCMs revealed broad increases in accessibility of inflammatory proteins FOSL2, JUN, JUNB, JUND, FOS, FOSL1, BATF and FOSB as well as several dimers of JUN and FOS family genes (Figure 9). These dimers are collectively called AP-1.(Vartanian et al., 2011; Zenz et al., 2008) Inflammatory mediators BACH1 and BACH2 also showed increased motif accessibility.(Hipp et al., 2017; Patsalos et al., 2019) MAITs only showed decreased accessibility of the ZBTB32 motif and CD8+ TCMs showed no motifs with significantly decreased accessibility.(Beaulieu et al., 2014) CD14+ monocytes had broad increases in accessibility of IRF motifs including IRF2/3/4/5/7/8/9 along with increased accessibility for the STAT1::STAT2 dimer. These cells also showed decreased accessibility of SOX4.

**Figure 9:**
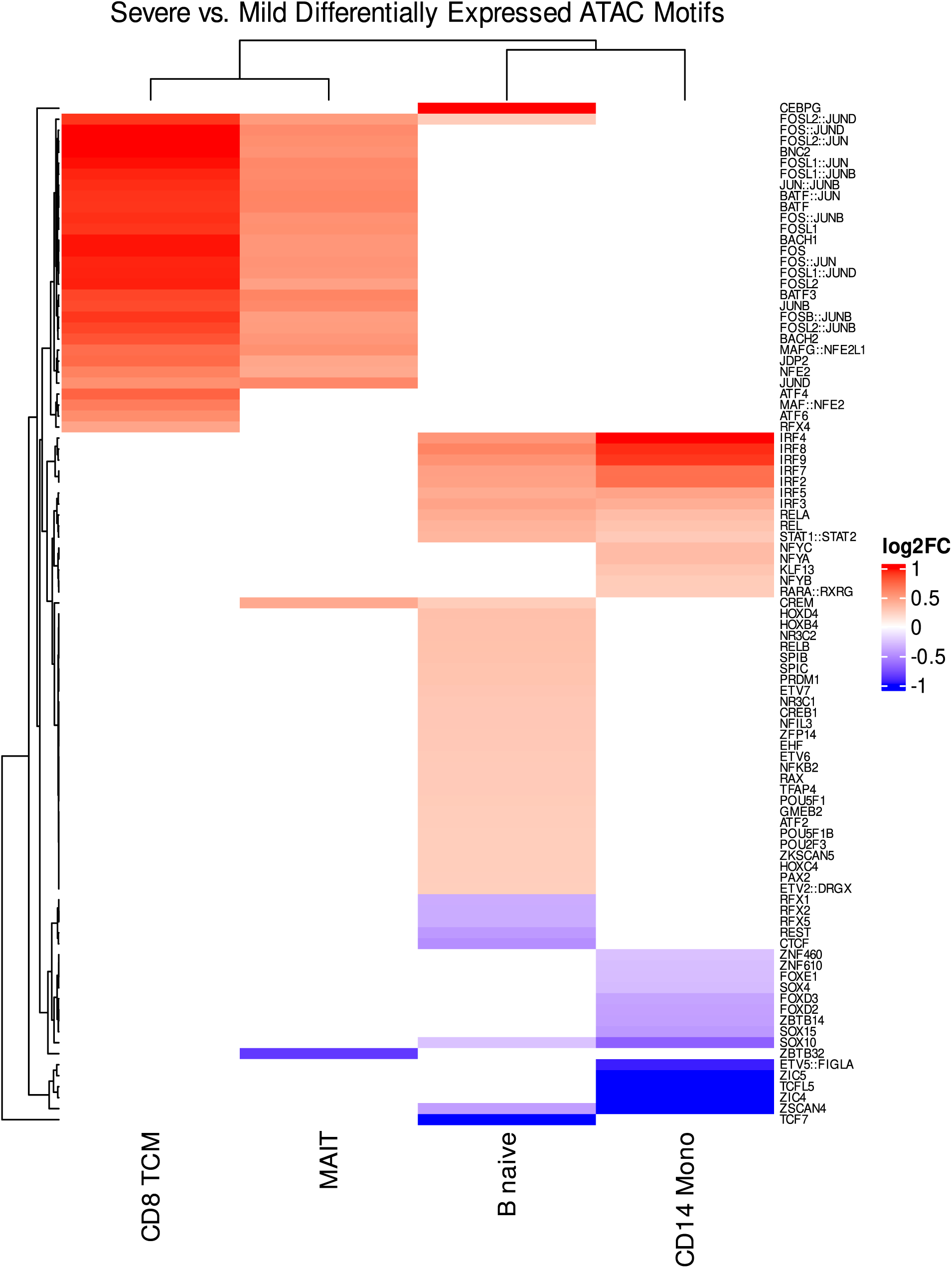
Differentially expressed motifs detected from ATAC-seq analysis of severe vs. mild cases. All detected motifs that were significant within at least one of the four top-ranked cell types are shown. Dimer motifs are indicated by “protein1::protein2”.

Naïve B cells also showed increased accessibility of the same IRF motifs along with the STAT1::STAT2 dimer motif. Additionally, increased accessibility of NR3C1, NR3C2, and NFKB2 was observed. The TCF7 motif was found to be decreased in accessibility.

Accessibility of ATAC peaks associated with genes showed low numbers of differentially accessible peaks for MAITs and CD8+ TCMs. Decreased accessibility of RUNX2 was found in both along with increased accessibility of HLA-C (Figure 10). KLF2 was also decreased in CD8+ TCMs but not in MAITs. CD14+ monocytes and naïve B cells had more differentially accessible peaks. CD14+ monocytes exhibited increased accessibility of HLA-C as well as decreased accessibility of many immune-related genes including CCL2, FOSL2, ELF2, NFKBIE, RUNX1, CCR3, KLF1, IL4R CXCR4 and BCL11A.

**Figure 10:**
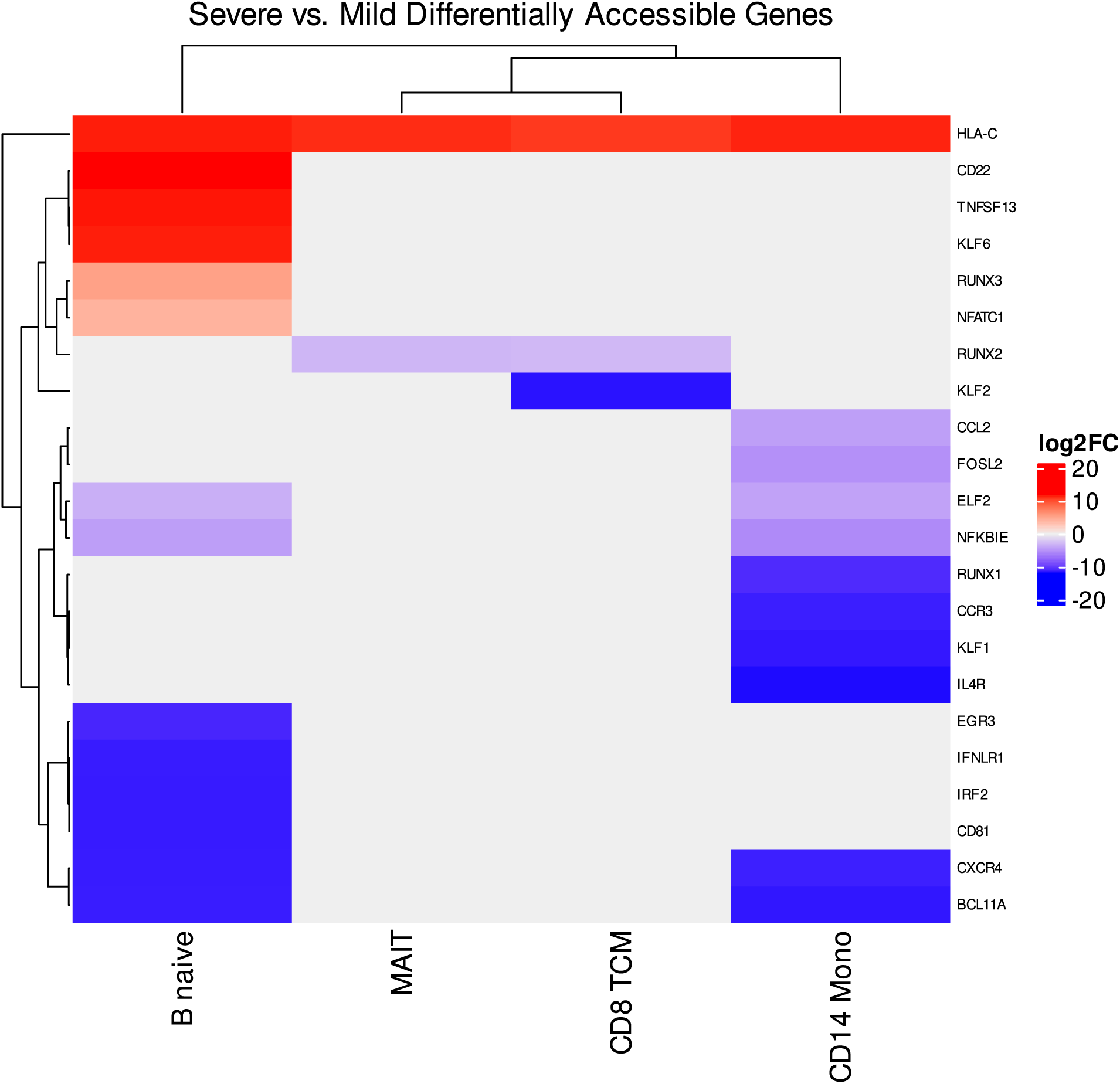
Differentially accessible genes detected via analysis of ATAC-seq data between severe and mild cases. Only genes of immunological significance are shown.

Naïve B cells also share increased accessibility of HLA-C. Additionally, these cells showed increased accessibility of genes involved in inflammation and B cell function regulation such as KLF6, TNFSF13, and CD22. These cells exhibited increased accessibility of NFATC1 which has a role in B cell activation and can both promote and suppress B regulatory cell activity through modulation of IL10 expression.(Catalán et al., 2021; Giampaolo et al., 2018; Li et al., 2018; Michée-Cospolite et al., 2022a) Naïve B cells also showed decreased accessibility to the B cell signaling genes CD81 and BCL11A. Additionally, decreased accessibility of interferon signaling genes such as IFNLR1 and IRF2 as well as CXCR4 were also observed.

### Severe Sarcoidosis is Characterized by Immunodysregulation Across Multiple Cell Types

We utilized single cell multiomic methodology to further interrogate differences in the immune environment of severe sarcoidosis versus mild sarcoidosis. We reveal cell-type level patterns that expand upon our bulk RNA-seq findings and provide a new framework for understanding severe sarcoidosis. Overall, our multi-omic analysis of sarcoidosis severity revealed various immunological pathways that differ between mild and severe disease. We successfully integrated our data between the 3’ and multiome kits, creating a shared set of cell identifications that enable reproducible analyses. The observed differences in cell abundance between the two kits are most likely due to the significantly harsher extraction protocol utilized in the multiome kit which involves more handling and cell lysis.

The combined analysis of DEGs showed GO term enrichment patterns that portray a complex mix of immune activation and suppression. This is consistent with the current knowledge of sarcoidosis and its juxtaposition of hyperinflammatory activity with anergy.(Miyara et al., 2006) Additionally, we find evidence of increased naïve B cell activation and Breg function which has been previously observed in sarcoidosis patient.(Saussine et al., 2012)

Closer inspection of the DEGs and DERs of sarcoidosis patients revealed more cell type specific patterns of immune dysregulation. Our top ranked cell types from differential analysis can be roughly split into two groups. The naïve B cells appear to be perturbed in one direction while CD56bright NKs, MAITs, CD8+ TCMS, CD14+ monocytes, and CD4+ TEMs show largely opposite patterns of activity, with CD14+ monocytes having distinct DEG patterns but not distinct DER patterns from the other cell types in the group.

By analyzing differential patterns across all cell types, we observed activity that could be grouped into four types: hyperinflammation drivers, inflammatory contributors, unresponsive, and alternative response. (Figure 11) The hyperinflammation drivers group is characterized by the CD56bright NKs, MAITs, CD8+ TCMs and CD4+ TEMs. This group exhibited broad activities of multiple immune response pathways throughout our analyses that are centered around AP-1 and its related signaling systems.(Vartanian et al., 2011) Concurrently, these cells also exhibited decreased activity of some immune pathway regulators that control such responses. This signifies that partial feedback responses may be suppressing portions of the immune regulatory system in these cells, but there is still a strong enough immune activation signal to continue pushing their proinflammatory activity.

**Figure 11:**
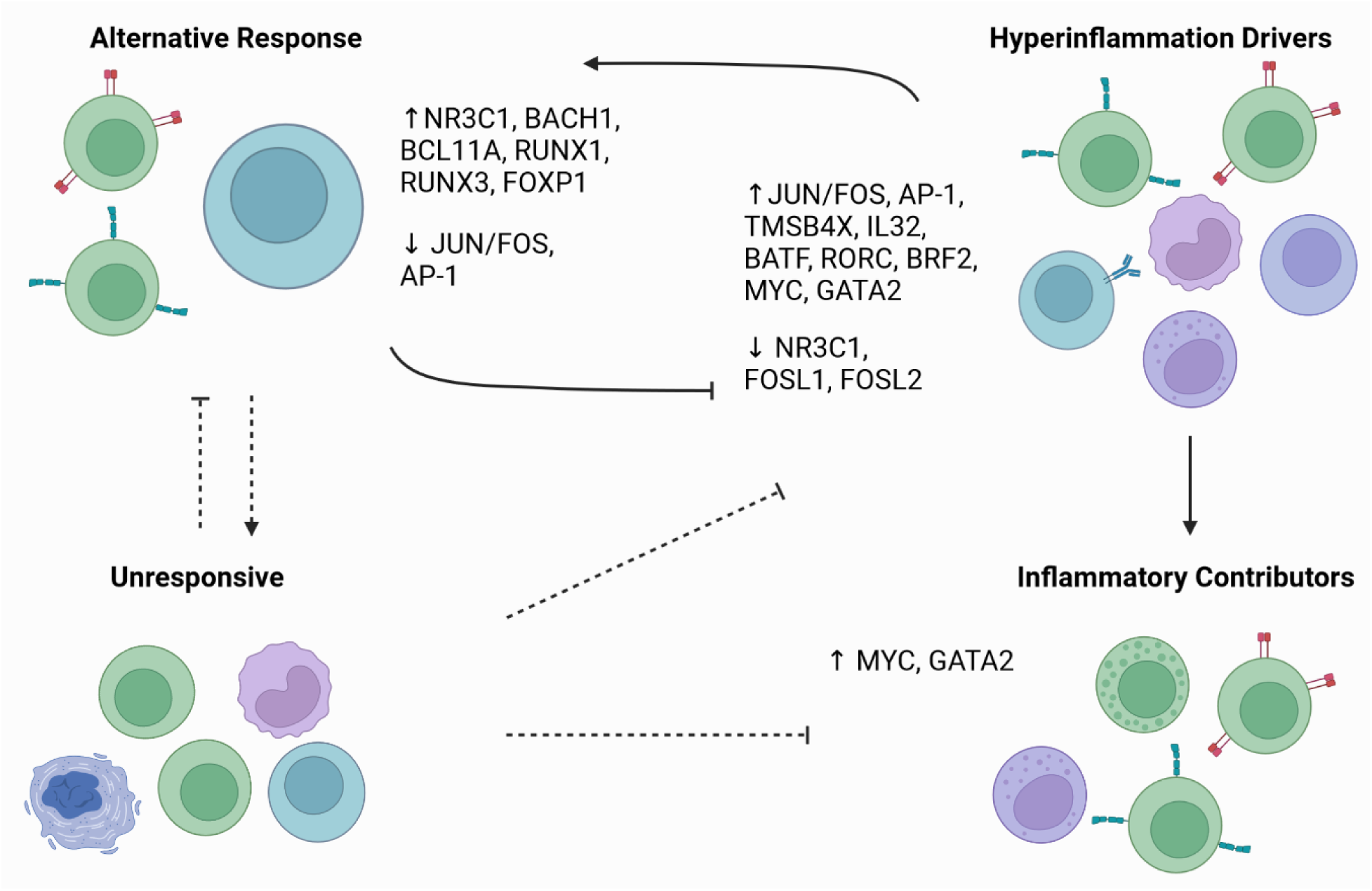
Summary of Expression Patterns in Severe vs Mild Sarcoidosis. The hyperinflammation drivers group is characterized by the CD56bright NKs, MAITs, CD8+ TCMs and CD4+ TEMs. The inflammatory contributors group consists of many more immune effector cell types including NKs, CD4+ TCMs, CD4+ CTLs, naïve CD8+ cells, memory B cells, CD14+ monocytes, and gdTs. The unresponsive group contains platelets, dnTs, Tregs, CD16+ monocytes and B intermediate cells. Lastly, the alternative response group is composed of naïve B cells, naïve CD4+ cells and CD8+ TEMs.

The inflammatory contributors group consists of other cell types that shared similarity to the hyperinflammation drivers group but had less dramatic differences between mild and severe cases. This group includes many more immune effector cell types including NKs, CD4+ TCMs, CD4+ CTLs, naïve CD8+ cells, memory B cells, CD14+ monocytes, and gdTs. The unresponsive group contains cell types that had minimal changes in activity between mild and severe disease. This included platelets, dnTs, Tregs, CD16+ monocytes and B intermediate cells. Strikingly, this group contains many of the cell types that are considered to have immunoregulatory activity including dnTs, Tregs, and CD16+ monocytes.(Kapellos et al., 2019; Wan, 2010; Wu et al., 2022) The lack of mobilization of these cells in severe patients stands in stark contrast to the disorganized activation and deactivation of various immune pathways seen in the hyperinflammation group. This highlights a core problem in sarcoidosis where immune regulation is not working in sync with immune activation.

Lastly, the alternative response group is composed of naïve B cells, naïve CD4+ cells and CD8+ TEMs. This group is particularly defined by the activity of naïve B cells which show patterns of immune activation that are distinct, and often directly opposed to the patterns observed in the hyperinflammation group. The increased activity of B cell activation associated genes and regulons in these cells is contrasted by decreased activity in other immune response systems, particularly those surrounding AP-1 activity. Furthermore, we find evidence of Breg activity through activation of FOXP1 as well as accessibility of NFATC1. Since IL10 expressing Bregs cooperate with Tregs in immunosuppression, and we observed minimal Treg changes, this finding may indicate a breakdown in signaling between Bregs and Tregs that may be related to dysfunctional IL10 signaling between the two cell types and poor Treg function.(Chaudhry et al., 2011; Michée-Cospolite et al., 2022b; Saussine et al., 2012)

The AP-1 pathway, which includes activation via the JUN family of transcription factors and mediates many downstream immune regulators including MYC and GATA2, seems particularly linked to both the hyperinflammatory group and the alternative response group. In the hyperinflammatory group, JUN family regulons and their downstream counterparts are heavily activated while the same group of regulons is suppressed in the alternative response group.

Our work is limited by the small number of individuals observed and the fact that they only represent one regional sarcoidosis population and one definition of severity in sarcoidosis. Additional work expanding on the size of our study and targeting some of the specific cell types we assessed may yield further insights into the mechanisms of the immunological disarray we observed.

Taken together, our findings indicate a new model of the transition to severe disease in sarcoidosis. Runaway immune activation driven by two opposing mechanisms may cause impaired feedback to immune regulator cells, hampering their ability to properly respond to the dysregulated immune system. This loss of immune cell communication appears to suppress or decouple Treg activity and provides insights into why sarcoidosis can be highly challenging to manage.

## Data Availability

All data produced will be made publicly available via GEO once the manuscript is accepted for publication.

